# Periodic intensification of routine immunization (PIRI): modeling a novel strategy to supplement routine and pulsed measles vaccination

**DOI:** 10.64898/2026.02.12.26346210

**Authors:** Kaiyue Zou, Matthew Ferrari

## Abstract

**Background:** Routine immunization (RI) is widely used to increase population immunity against measles. In low-resource settings, achieving immunity goals using RI alone has proved challenging and supplemental immunization activities (SIAs), large community-based vaccination campaigns conducted every few years, have been used to close immunity gaps. Although effective at covering the population unreached by RI and boosting the population immunity, SIAs are labor-intensive and expensive, allowing for accumulation of susceptible in-between campaigns. Periodic intensification of routine immunization (PIRI) has been used in settings with high measles incidence as another supplement to the existing immunization framework. PIRI is built on the existing RI infrastructure and often more frequently than SIAs. However, the effects of PIRI have not been quantitatively evaluated against SIAs.

**Methods:** We developed a stochastic age-structured SIR model parameterized by measles dynamics to simulate RI, RI plus SIAs, and RI plus PIRI vaccination strategies to quantify their effects on measles control. We define the sufficient PIRI rate as the immunization rate during a PIRI, relative to RI, that achieves equal or fewer cases than a corresponding RI plus SIAs strategy.

**Results:** We found a U-shaped relation between the sufficient PIRI rate and RI coverage levels. When RI coverage levels are low or high, PIRI cannot reduce cases below the level achieved by RI plus SIA. But at intermediate RI coverage levels, the sufficient PIRI rate is low; approximated 4-times the RI rate, and PIRI achieves fewer cases than SIAs. We also found that in scenarios of increasing RI coverage, maintaining PIRIs even after this intermediate regime results in a negligible increase in cases and lower annual variability.

**Conclusion:** PIRI has the potential to meet or exceed the performance of an SIAs-based strategy in settings with intermediate levels of RI coverage or settings with high and continuously increasing RI coverage.

## Introduction

Vaccination against measles has averted 60 million childhood deaths since 2000 ^1^. In low and lower middle-income countries, vaccination against measles has conventionally been delivered through a combination of routine immunization (RI) and supplemental immunization activities (SIAs). RI is carried out regularly at fixed health service centers through well-child visits at targeted age and is (intended to be) recorded in the child’s vaccination history ^2,3^. SIAs are occasional, pulsed vaccination campaigns that target all children within a wide age range and are delivered through community-based sites that are designed to reach children that are not in regular contact with the health service centers that provide RI service. Each mechanism has limitations but they work in concert to maintain high population immunity ^4,5^.

Achieving and maintaining high RI coverage comes with great challenges as it relies on the accessibility of vaccination centers. Low trust or awareness of RI services among caregivers, transportation barriers, deficiency in trained health workers, lack of precise planning and logistics management in vaccination, and limited number of RI sessions ^3,6^ have all been linked to low coverage of the RI system. RI coverage for measles had been increasing before 2019 but plateaued at ∼85% globally during the 2015 to 2019 period ^6^. Correspondingly, measles cases have increased globally over the last 6 years^7^.

WHO recommends immunization coverage be above 95% with two doses to eliminate measles ^8^. Even with sufficient resources, there may be significant operational challenges to achieving this high coverage in under-resourced settings. Lack of timely and accurate demographic data may limit the planning and the effectiveness of RI ^9,10^. For instance, polio eradication activities and its RI planning in Nigeria relied on census data with a 3% projected birth rate although the real population growth was faster than that ^11^.

Supplemental Immunization Activities (SIAs), or catch-up SIAs specifically ^12^, are designed to provide a first-dose opportunity to those missed by the routine program and to provide a second-dose opportunity where routine second-dose coverage is low ^6^. With the goal of complementing RI ^13^, SIAs are resource-intense, large-scale vaccination campaigns ^3^ to immunize a large number of children. Unlike RI, SIAs are conducted every 2 to 5 years ^5,14^ and aim to cover a wide age range ^3^ regardless of the child’s vaccination status ^5^ (i.e. they are non-selective with respect to prior vaccination). To increase reach into unvaccinated communities, SIAs deliver vaccines outside of conventional health facilities and engage societal entities (e.g. community leaders and political leaders) to encourage uptake ^3^.

Doses delivered via SIAs are not included for the annual vaccination coverage estimation^3^. Multiple studies have shown that SIAs’ effectiveness at reaching “zero-dose” kids, i.e., the population missed by RI, is variable; a study focused on low- and middle-income countries found that measles SIAs accomplished a 66% coverage, on average, among children that received no prior measles vaccines across the countries ^5^. A study conducted in India reported measles-rubella SIAs reached between 45.8% to 94.9% of “zero-dose” kids ^15^. A study in Zambia reported 27.8% of the “zero-dose” kids were covered by measles-rubella SIAs, decreasing the proportion of “zero-dose” kids from 15.1% to 10.9% ^16^.

Despite the demonstrated benefits of SIAs, they remain expensive to implement ^17^, and the non-selective nature means they inevitably vaccinate many immunized children instead of only reaching those at risk. This redundancy necessarily increases with increasing population immunity. High coverage of RI may make SIAs less efficient and less cost-effective, per dose. Narrow age range SIAs (e.g. under 5 years) may miss many unimmunized people as the RI coverage increases ^18^. A study in Benin found lower cost-effectiveness of SIAs is associated with higher coverage of RI ^19^. Also, in practice, reaching unvaccinated kids becomes more difficult with higher RI coverage ^20^. On the other hand, the cost of SIAs may disproportionately burden weaker health systems^21^. The long time between SIAs may fail to prevent outbreaks if the RI coverage is not sufficiently high ^6,12,22^, particularly if SIAs are delayed ^7^. Finally, countries with low coverage of RI are more reliant on frequent SIAs ^6^ and this dependence may divert resources necessary for improving delivery via RI ^12,23^.

Periodic Intensification of Routine Immunization (PIRI) was first formally documented as an immunization program by WHO in a preprint in 2009 ^24^. PIRI provides RI services, that are selective of un- or under-vaccinated children^25,26^, over short periods ^26^, analogous to SIAs ^25^. PIRIs are usually combined with other child and maternal health services ^24^. PIRI examples include “Vaccination Week” in the Americas and “European Immunization Week” that are conducted in countries with higher RI coverage ^24^, Child Health Days (CHD) and Immunization Weeks (IW) and Intensified Mission Indradhanush (IMI) in India ^25,27^, and Child Health Days and African Vaccination Week (AVW) in Africa ^28,29^. PIRIs are usually more frequent (one per year or more than one per year) ^24,28^ than SIAs, providing more opportunities to reach children missed by RI ^6^. Unlike SIAs, PIRIs are disproportionately implemented through the network of facilities that deliver RI, and thus can potentially have secondary benefits on RI infrastructure in countries with weaker health systems ^24^. Unlike SIAs, doses delivered via PIRIs are recorded as part of the vaccination history ^25,26^.

A study done in India using data from 2007 to 2008 found that PIRI is effective in increasing the RI coverage and immunizing kids who would be missed by RI services, especially in areas commonly considered hard to reach by RI ^25^. A 2015-2017 national PIRI in India was found to be effective at increasing full immunization coverage among kids under 2 ^30^. A more recent study in India found that the 2017-2018 nationwide PIRI increased the diphtheria tetanus-pertussis (DTP) vaccination coverage among the unvaccinated kids by 35.7%; it’s also found this program increased the overall immunization coverage among kids by a range of 12% to 31% across different districts and multiple vaccines ^27,31^. Notably, 63% of the total vaccine doses in this PIRI program were delivered to the unvaccinated kids ^26^. The 2017-2018 PIRI cost twice as much as RI per dose, but was still estimated to be cost-effective in India compared to per-capita gross domestic product (GDP) per disability-adjusted life year (DALY) averted ^26^. A study in Cameroon found that PIRI added 4.5 USD to the vaccination cost per person compared to RI, but improved RI coverage and accelerated introduction of new vaccines during a period of armed conflict and the COVID-19 pandemic ^32^. Another study conducted in multiple African countries found that PIRI increased measles immunization coverage by 10%, with higher impacts in areas with lower RI coverage ^28^.

PIRI, though proven effective at improving RI in some cross-sectional studies, has not been formally and mathematically defined; this limits the use of mathematical models to evaluate of the potential for PIRIs to supplement RI and/or replace SIAs for measles control and elimination. Here we present a mathematical representation of PIRI within an age-stratified Susceptible-Infected-Recovered (SIR) compartmental modeling framework. We then use this model to compare the effectiveness of disease control in three different immunization scenarios: RI only, RI plus SIAs, and RI plus PIRI.

## Method

We used an age-stratified SIR compartmental model with maternal immunity (M) and vaccination-induced protection (P) to simulate vaccination via routine immunization (though a first and second dose), SIAs, or PIRI. The model was simulated with a discrete time-step of one day via the tau-leaping algorithm ^33^. We model 136 age groups in total, where individuals first pass through 60 month-long age groups (from 0 month old to 59 months old) and then 76 year-long age groups (from 5 years old to 80 years old and above). Without losing precision or the generalization of the study results, all months are assumed to be 30 days consistently and all years are assumed to be 360-day-long for the convenience of the aging process. The natural aging process is simulated in a pulsed manner ^34^: on the first day of each year, all age groups and month groups are moved upward; on the first day of each month, only the month groups are moved upward. At the beginning of each distinct day of simulation, the above aging process takes place first; then, if applicable, the vaccination via SIAs is conducted by directly and instantaneously moving a certain proportion, which is equal to the coverage (*ρ*) discounted by effectiveness (e), from the corresponding age-specific S to R compartment. Next, birth, death, maternal protection waning, infection, recovery, immunization through routine vaccination, or PIRI (if applies) processes are simulated simultaneously (Figure 1). Since these competing processes take place at their own fixed rates, there are cases where the total number of people leaving a compartment surpasses the current total number of people in compartment combination; in this event, the numbers of people leaving due to all sources are based on current total number of people in the compartment, normalized by the corresponding rate relative to the rates sum.

**Figure 1.**
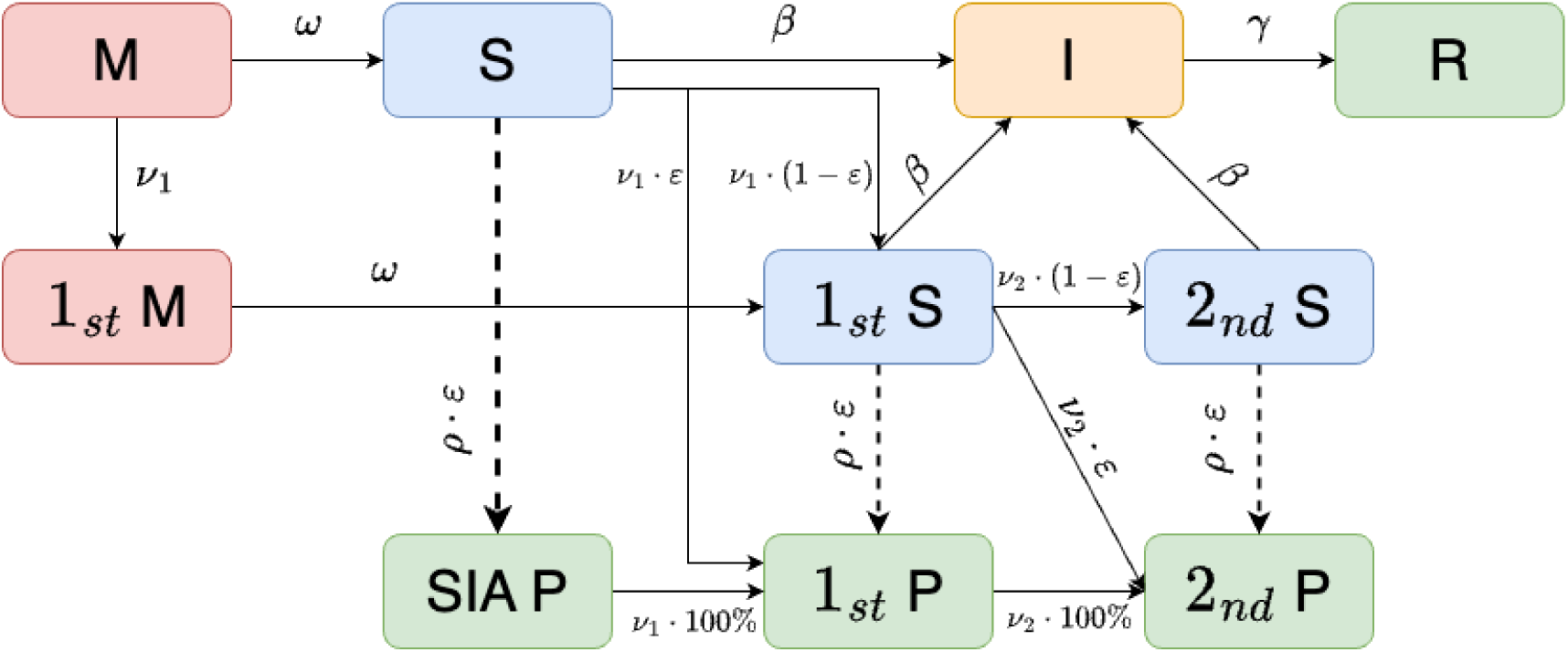
The flowchart of the age-structured SIR model. Class M represents individuals with maternal immunity. Class S represents susceptible individuals. Class I represents infected individuals who are also infectious. Class R presents individuals who recovered from infection and gained lifetime immunity. Class P represents individuals who are immunized from either RI, PIRI, or SIAs. “1st” and “2nd” mark how many doses of RI or PIRI vaccines the individuals have received, which are assumed to be recorded in their vaccination history and are retrievable. In short, Class P and Class S indicate the immunity status; “1st”, “2nd”, and the absence of both indicate the retrievable vaccination history. Class SIA-P represents individuals who are fully immunized by SIAs but their vaccinations through SIAs are not recorded and not retrievable. Solid lines represent processes happen at a specific rate (associated with RI and PIRI); dashed lines represent processes happen by moving a specific proportion of individuals from one class to another (associated with SIAs). Parameter definitions and values can be found in Table 1.

**Table 1.** Parameters used in simulations that have fixed or bounded values. Not all the parameters from the simulations are included above: for example, age-specific 1st dose RI vaccination rates 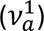 and age-specific PIRI vaccination rates 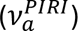 highly dependent on simulations; PIRI target age range also differs across simulations.

| Parameter Definition | Symbol | Value or Range |
| --- | --- | --- |
| Initial total population size | $N$ | 961,000 |
| Initial infectious age group |  | 5 months old |
| Initial infectious person(s) |  | 10 |
| Initial susceptible proportion among 14 months old and above |  | 10% |
| Initial susceptible proportion age range lower bound |  | 13 months old |
| Annual birth rate for the reproductive age groups | $\lambda$ | 50.0/1000.0 |
| Reproductive age range |  | [18, 50] |
| Mean age of life | $1/\mu$ | 80 years |
| Basic reproduction number | $R_0$ | 16.0 |
| Recovery rate | $\gamma$ | 1.0/8.0 |
| Rate of infection | $\beta$ | 2.08174e-06 |
| Seasonal multiplier on the rate of infection |  | [0.5, 1.5] |
| Mean duration of maternal immunity | $1/\omega$ | 6 months |
| Measles seroconversion rate per vaccination | $\varepsilon$ | 95% |
| Coverage of the first-dose routine vaccine | $P\%$ | [0%, 100%) |
| Lower age bound of the first-dose routine vaccine |  | 6 |
| Peak age of the first-dose routine vaccine |  | 9 |
| Upper age bound of the first-dose routine vaccine |  | 14 |
| Coverage of the second-dose routine vaccine (fixed) |  | 85% |
| Lower age bound of the second-dose routine vaccine |  | 15 |
| Peak age of the second-dose routine vaccine |  | 18 |
| Upper age bound of the second-dose routine vaccine |  | 24 |
| SIAs coverage | $\rho$ | 70% |
| SIAs frequency |  | Every 5 years |
| SIAs duration |  | 1 day |
| SIAs target age range |  | 9 months old to 59 months old |

The basic reproduction number (R0) is 16 ^35^ and remains constant across all scenarios. The default rate of infection is determined by the initial overall population size *N*, the overall death rate *μ*, and recovery rate *γ* using the formula: 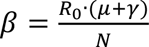. Measles seasonality, which effectively is a scalar applied to the rate of infection dependent on the time of the year, is assumed to be a shifted sine function of time in the year and peaks in June annually. Each age group has an age-specific initial population size (STable 1), birth rate (STable 2), and death rate (STable 3), the combination of which roughly follow a pattern common in developing countries with growing population size. For example, the population size decreases monotonically from younger age group to the older ones. We assumed that the mean length of period of infection is 8 days and the mean length of period of maternal protection is 6 months. We assumed homogeneous mixing pattern among all age groups. The results from heterogeneous mixing are available in the Supplement. We assumed commuter-style case imports to prevent the local extinction of transmission, where the susceptible people within the population will become infected at a daily number quantified by formula: 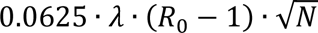, where *λ* represents the birth rate and *N* represents the total number of people ^36^.

The RI process is simulated continuously across all scenarios. First-dose RI and second-dose RI vaccine are administered at rate 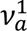 and 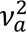, respectively, for a given age *α*. The age-specific RI rate 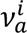 is assumed to be proportional to a normal distribution with mean (i.e. peak vaccination rate) at age 9 months (for the first dose) and 18 months (for the second dose); the standard deviation controls the degree of early and late vaccination though RI and is assumed to be 1.5 months for both routine doses (Figure 2), i.e., 68% of the first-dose RI vaccines are administered to kids between 7.5 and 10.5 months old. Only those who received the first dose of vaccine with a retrievable record, either from RI or PIRI, are eligible for the second dose of vaccine. Dependent on the vaccine efficacy, e, the susceptible people, once vaccinated, either enter the protected class or another susceptible class with an updated vaccination status (e.g., a failed 2nd dose RI vaccine will convert 1st Susceptible to 2nd Susceptible).

**Figure 2.**
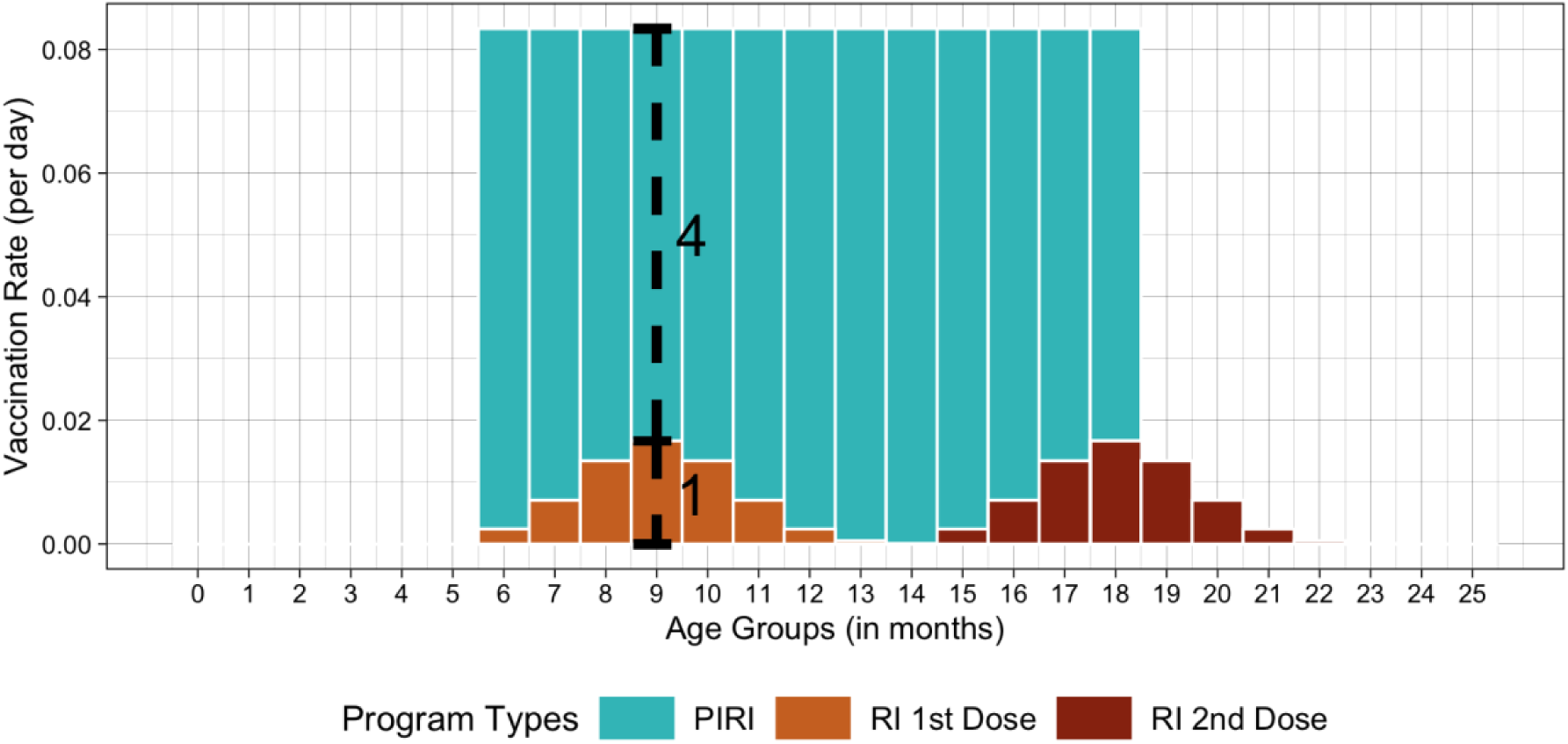
The illustration of vaccination rates as a function of age for the RI and PIRI programs. This figure is specific to the following setting: the first-dose RI covers 85% of the kids who grow out of 14 months old, the second-dose RI covers 85% of the kids who have received their first dose RI vaccine and grow out of 24 months old, and the ratio between the PIRI rate and peak rate of first-dose RI is 5.

Vaccination via PIRI is simulated periodically (here, annually) beginning and ending on a specific date (here, PIRI lasts throughout March each year if not stated otherwise). During the PIRI month, the age-specific vaccination rates of the first-dose RI (which is usually the case unless stated otherwise), the second-dose RI, or the both of them are set to 0; RI effectively occurs through PIRI, utilizing the PIRI rates. In general, the PIRI vaccination rates occur through a new age-specific vaccination rate 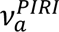; specifically, we model 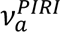 as constant, *C*, for all ages *α* between *A*^*lower*^ and *A*^*upper*^. We assume that *C* is always greater than the maximum of 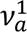 and define the “PIRI-to-RI rate ratio” as 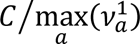 (shown in Figure 2, where the ratio is 5). We choose this metric to define the ratio between PIRI and RI instead of ratio between the overall PIRI rate and overall RI rate integrated across covered age groups 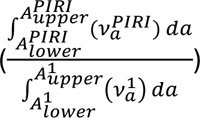 for several reasons. RI is normally recommended for a narrow targe age range; in reality, however, the variance in the distribution of vaccination rates over age groups becomes an emergent property instead of an operational intent, i.e., age groups outside the target also get RI vaccines in practice. This phenomenon has been captured across countries and times in a study by Timos Papadopoulos et al ^37^. We’ve chosen to use the vaccination rate associated with the recommended RI target age group (or the peak rate of RI) because that’s an operational age within the realm of vaccination program designs and with epidemiological significance. This metric also remains easier to understand when we expand the target age range of PIRI to be wider or loosen the selectivity of PIRI.

Nevertheless, because of the variance in the vaccination rates distribution, it is useful to consider the ratio between two integrated rates since it emerges to be the realized rate ratio between two vaccination programs in reality. Therefore, and hereafter, we call 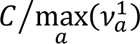 the rate ratio between PIRI and RI, or the PIRI-to-RI rate ratio; and we will present the results using the other metric in the Supplement (SFigure 3 and SFigure 4).

Vaccination via SIAs is simulated every 5 years by default; it’s assumed to happen and conclude over the first day of the 3rd month of the SIAs year. Unlike RI or PIRI, the SIAs are not recorded in the vaccination history, and a proportion equal to the coverage (*ρ*) times the effectiveness (*ϵ*) will move from the susceptible class to the corresponding protected class for all eligible age classes without changing the on-record vaccination status. Thus, individuals receiving their first or second dose via SIAs are still eligible for the first or second dose, respectively, from either RI or PIRI since they’ve received zero or only one dose of vaccine based on the record. We assumed SIAs achieves coverage of 70% among anyone between 9 months and 59 months old.

For vaccination mechanisms, we assumed a homogeneous seroconversion rate, *ε*, at 95% for any given dose of vaccine; in practice this will differ by age at administration ^38^, but will not qualitatively impact the results here. We assumed the rate of first-dose routine vaccination has peak vaccination rate for the 9-month-old age group with *P*% of vaccinated individuals receiving doses between 6 months old to 14 months old. We varied the magnitude of the age specific vaccination rate 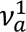 such that *P*% of individuals were vaccinated by age 14 months, where *P*% is the vaccination coverage of the first dose in the RI system in a given scenario. We assumed the rate of second-dose routine vaccination peaks in the age group of 18 months with a fixed 85% coverage among age groups between 15 months and 24 months. Notably, we also assumed the second-dose routine vaccines are only distributed among those received their first dose. We then calculated the age-specific vaccination rates 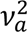 for the second-dose RI such that 85% of individuals among those received their first dose were vaccinated by the age of 24 months.

We simulate three vaccination scenarios: RI only, RI plus SIAs, and RI plus PIRI. Across all three scenarios, we hold the demographic and transmission parameters constant and only vary the characteristics of the vaccination sources. Across simulations within scenarios, we vary the coverage of the 1st dose of RI (from 0% to 100%) inside the RI only and RI plus SIAs scenarios; we vary the combinations of the PIRI rates (relative to 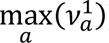) and the coverage of 1st-dose RI inside the RI plus PIRI scenario. For a singular simulation within each vaccination scenario, we simulated a time series of 10,000 days with constant vaccination characteristics (e.g., vaccination rates, target age groups, etc.). We then quantified the total number of cases over a period from day 5,001 to day 10,000 in each simulation. For each given level of RI coverage, we scanned across all the simulations in the RI plus PIRI scenario to identify the sufficient PIRI rates that resulted in the same or fewer cases than the corresponding RI plus SIAs scenario; the minimum of the identified sufficient rates is the minimally sufficient PIRI rate required for the RI plus PIRI scenario to achieve equal cases as the RI plus SIAs scenario—any PIRI rates higher than it will result in fewer cases in the RI plus PIRI scenario. The minimally sufficient PIRI vaccination rate over the peak of the RI vaccination rate using the formula 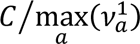 then becomes the minimally sufficient rate ratio, or MSRR, which will be measured across all RI coverage levels. Upon completion of the initial search for MSRR, we varied PIRI’s selectivity on children’s vaccination history, PIRI’s target age range, PIRI month of the year, and birth rate to generate more results.

To explore the transient impact of PIRIs in the real-world scenario where the RI coverage improves overtime, we also simulated the model in 6 scenarios with dynamical vaccination designs (which are described in Table 2) over 33,000 days with temporally changing RI coverage levels. We first simulated dynamics for 18,000 days in the absence of vaccination and then introduced RI that increased linearly from 0 to 100% over a period of 15,000 days (2.4% increase per year). Relative to the U-shape curve of the MSRR (described in Results), this gives three distinct stages: in Stage 1, RI is below the level for which SIAs result in fewer cases than PIRIs; in Stage 2, RI is in the range where PIRIs can result in fewer cases than SIAs if above the MSRR; and in Stage 3, RI is high enough that SIAs (because SIAs are non-selective) results in fewer cases than PIRIs. We simulated 6 scenarios including a baseline RI only scenario, RI + SIAs (where SIAs target ages from 9 months old to 5 years old, with coverage 70%, every 5 years), and 4 scenarios that either use PIRI (where PIRIs target ages 6 months old to 14 months old every 1 years) at all times, or switch from SIAs to PIRIs to SIAs in stages 1, 2, and 3 respectively. We assume that PIRI achieves either a constant vaccination rate ratio of 7 relative to RI, or the MSRR. The relative rate 7 was chosen arbitrarily to reflect daily vaccination consistent with weekly RI vaccination. For each scenario, we present the average cumulative number cases over time for 500 stochastic runs.

**Table 2.**
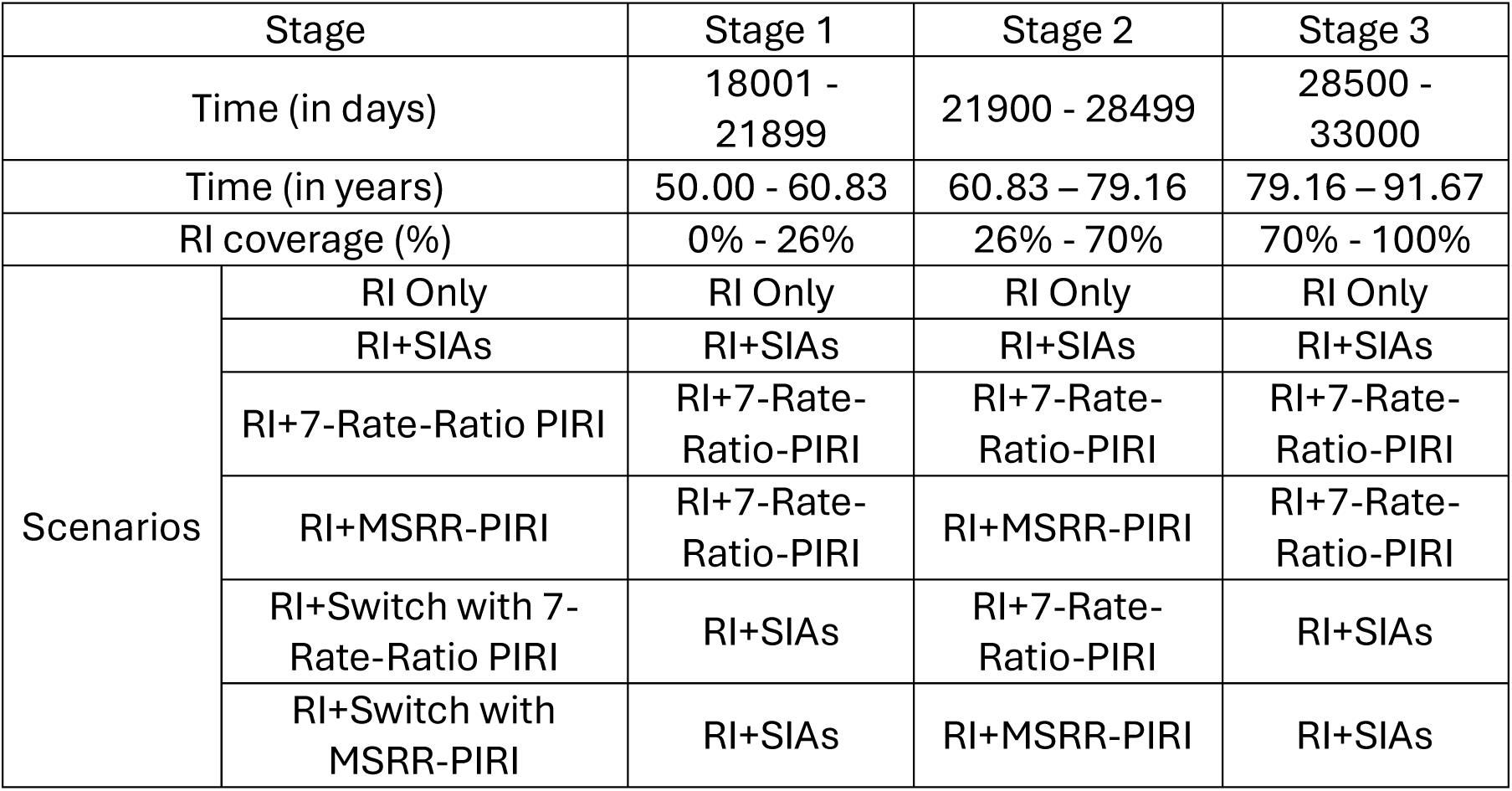
Vaccination program designs over different scenarios and different stages in. **Figure 5**.

## Results

The minimally sufficient rate ratio between PIRI and RI required for PIRI strategies to have equal cases as SIAs, or MSRR, is a U-shaped curve across levels of RI coverage from low to high (Figure 3). The area above the MSRR curve represents PIRI-to-RI rate ratios that result in fewer cases for PIRI than SIAs strategies. As illustrated below (in Figure 4 and in the Supplement), the explicit position of the MSRR curve is a function of various characteristics of the PIRIs and SIAs, as well as the demographics of the population; the general shape is robust. For the set of conditions illustrated in Figure 3, no rate ratio is sufficient to out-perform SIAs based strategies when RI coverage is below 10% or greater than 75%; outside these bounds, the SIAs strategy always leads to fewer cases than PIRI regardless of the rate of PIRI. Note that when RI coverage is low, so then is the maximal age-specific RI rate (Figure 2). Thus, for PIRI to make up for this low rate, the rate (during the short period of a PIRI) must be much higher. When the RI coverage is very high, on the other hand, most children will have been reached by RI and the population remaining susceptible is increasingly composed of those who were vaccinated but failed to seroconvert (primary vaccine failure). Since PIRI’s are defined to be selective of children that did not receive RI (this is relaxed below) and SIAs are non-selective with respect to past vaccination, the latter strategy performs better at high routine coverage levels. Focusing on the settings where RI coverage is intermediate (e.g. 25-70% in Figure 3), the MSRR is generally under 7 and can be as low as around 4.5.

**Figure 3.**
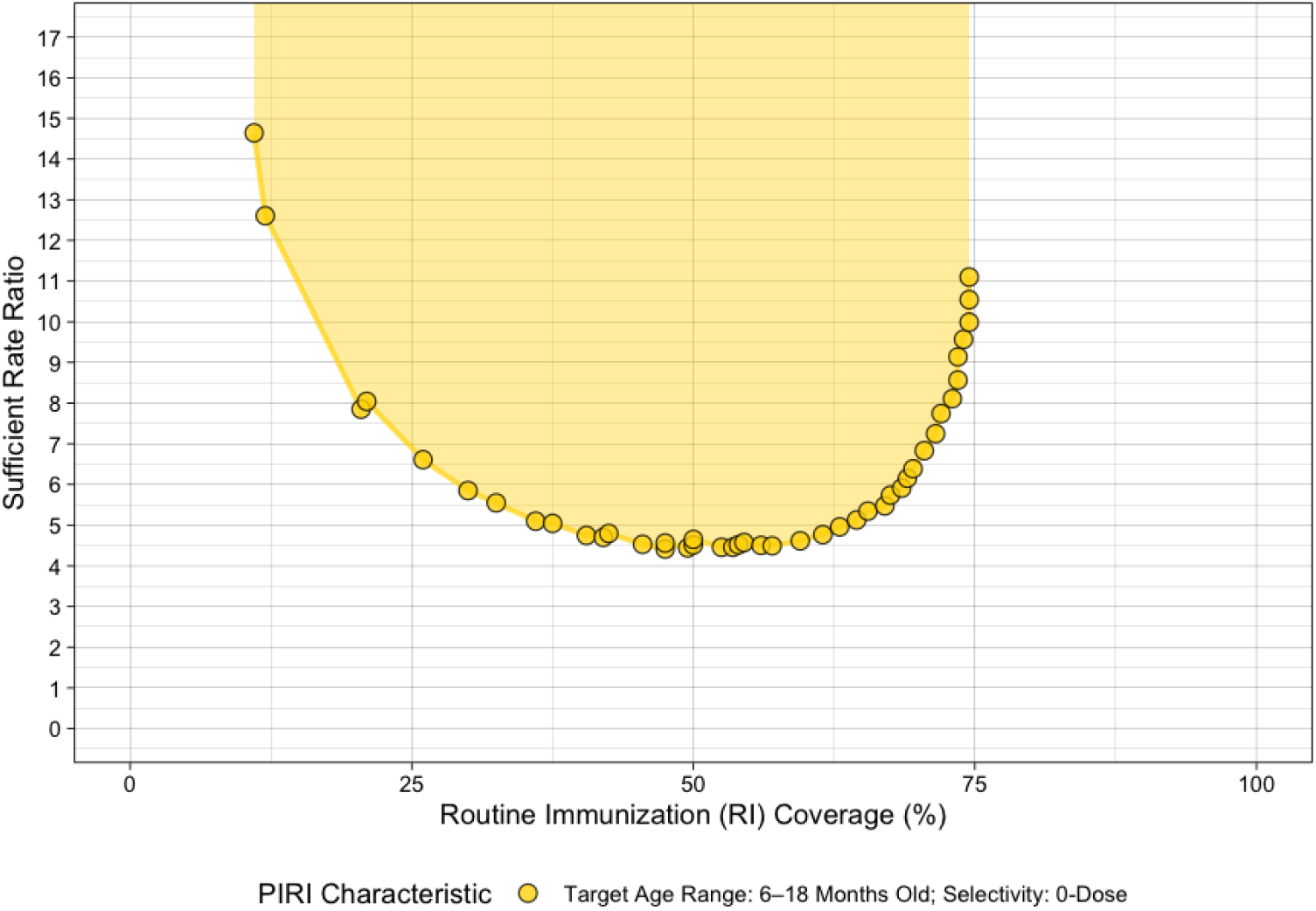
The sufficient rate ratio (shaded area above the curve) between PIRI and RI to achieve equivalent or fewer cases than SIAs at different levels of RI coverage during the period of stable dynamics (from day 5001 to day 10,000). PIRI targets kids 6 to 18 months old who received zero dose of vaccine from RI or PIRI before. The RI coverage levels vary from one simulation to another, but it remains unchanged within each scenario. SIAs always have a fixed 5-year interval and a 70% coverage amongst all kids under 5 years old across all simulations. PIRI’s vaccination rate remains fixed throughout each simulation but differs across different simulations.

**Figure 4.**
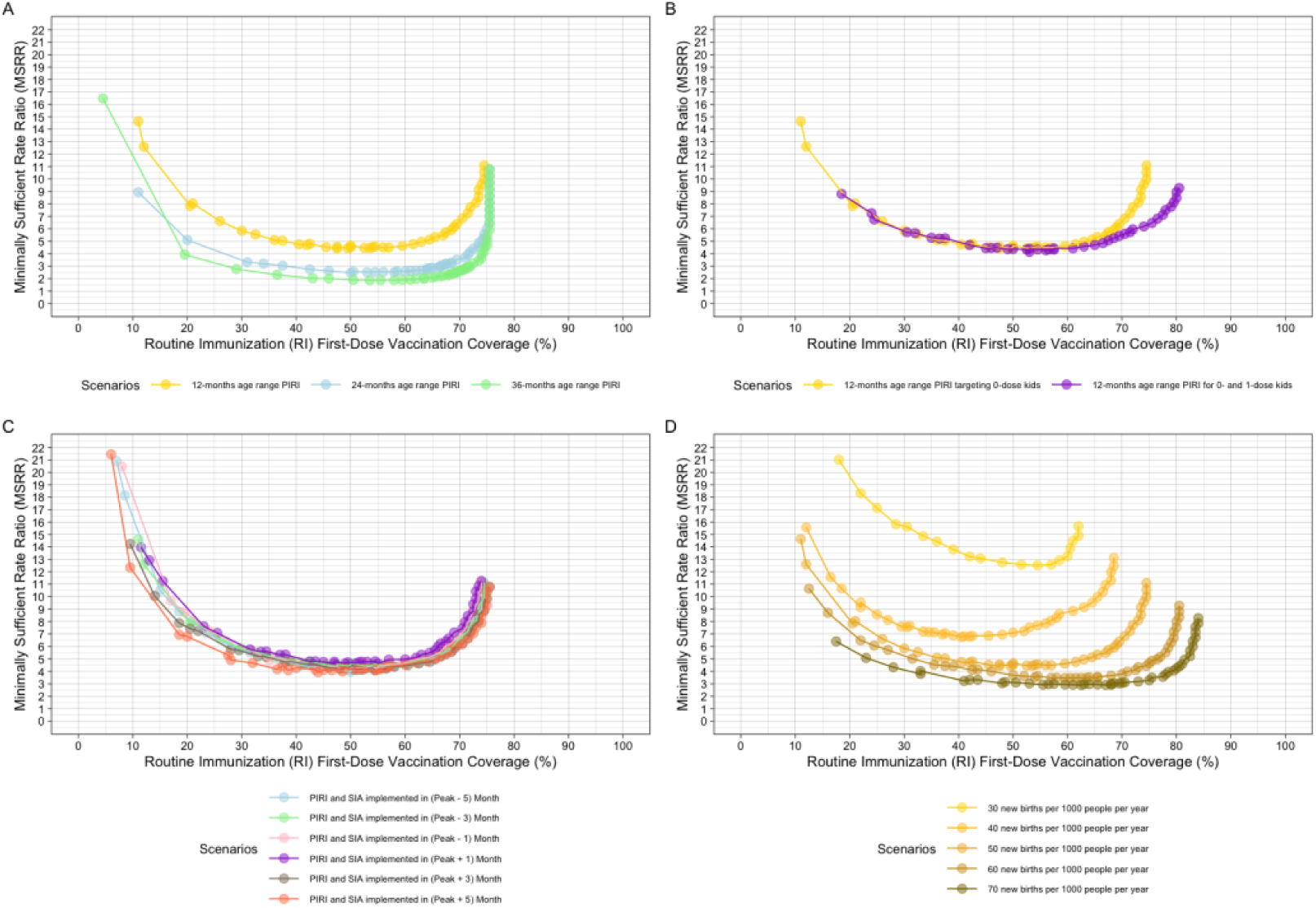
The minimally sufficient rate ratio (MSRR) across different levels of RI coverage under different assumptions of PIRI characteristics, SIAs characteristics, and demographic characteristics. Panel A demonstrates the MSRR curves under PIRIs with different ranges of target age group, from 12-month-long (the default, same as in Figure 3) to 36-month-long. Panel B shows the MSRR curve when PIRI targets only 0-dose kids (default) or 0- and 1-dose kids. Panel C compares MSRR curves when both PIRI and SIAs are implemented in a specific month relative to the peak month of the measles transmission, with the default scenario as the one where both SIAs and PIRI are implemented 3 months ahead of the peak month. Panel D illustrates MSRR curves given different birth rates in the population, from 30 new births per 1000 people at reproductive ages per year to 70 with the 50 as the default.

While the general U-shape of the MSRR curve is robust, the absolute position and the behavior at high RI coverage is sensitive to characteristics of the PIRI, the SIAs strategies to which PIRIs are being compared, and demographics of the population (Figure 4). The MSRR curve shifts closer to the x-axis, i.e. the minimum sufficient daily rate at which the PIRI vaccinates new individuals relative to RI decreases, as the age range of children eligible for the PIRI increases (Figure 4A). The marginal reduction in MSRR decreases as the age range increases from 24 months to 36 months (Figure 4A) when the SIAs interval is 5 years. Note that MSRR increases sharply at high RI regardless of PIRI age target (Figure 4A); as RI coverage increases there are fewer zero-dose children to be immunized through a PIRI, so the MSRR must be dramatically higher to provide marginal benefit.

Changing the selectivity of PIRIs, to allow vaccination of children with 0 or 1 dose, widens the RI coverage range over which the MSRR can remain relatively low (e.g. 10 or less in Figure 4B). For the settings illustrated here, there is no difference between a highly selective (targeting 0-dose children only) and less selective (targeting 0 or 1-dose children) PIRI over most levels of RI (RI < 60% in Figure 4B). Thus, a switch to less selective PIRIs, which would necessarily increase costs as more children are eligible, only yields net benefits as RI increases.

There are small differences in the height of the MSRR curve when PIRIs and SIAs are conducted before or after the seasonal peak of cases (Figure 4C). Consistent with previous studies on the timing of SIAs ^39,40^, MSRR is lowest (i.e. PIRI exceeds SIAs at lower rates) when administered out of phase with the seasonal peak of cases. In the Supplement we illustrate the MSRR curve relative to SIAs implemented at different frequencies or coverage (SFigure 1).

Increasing birth rates decreases the overall magnitude of the MSRR over all levels of RI and disproportionately for high levels of RI (Figure 4D). When birth rates are higher, PIRIs out-perform SIAs for a much lower set of daily relative vaccination rates and the rates are non-infinite for a wider range of higher RI coverage. This result occurs because high birth rates mean faster accumulation of the susceptible, and the annual cadence of PIRIs prevents susceptible accumulation that could allow large outbreaks in the wider intervals between SIAs.

We also derived the results and figures using an alternative metric defining rate ratio as the rate of PIRI integrated across covered age range divided by the rate of RI integrated across covered age range 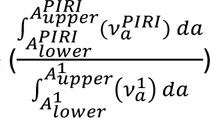, and they can be found the supplemental materials (SFigure 3 and SFigure 4). The results for this metric are qualitatively similar, though the correlation between PIRI age range and MSRR (Figure 4A vs. SFigure 4A) are reversed because the limits of integration in the numerator increase with the PIRI target age range.

To illustrate the impact of PIRI and SIAs strategies as RI programs increase over time, we simulated total cases under a simplified scenario with linear increase in RI over time, i.e., the x-axis in Figure 3. We compared scenarios that employed an RI only strategy, an RI plus SIAs strategy, two RI plus PIRI strategies, or two strategies that “switch” between SIAs when MSRR is undefined and PIRI when MSRR is defined and finite (intermediate RI) (Table 2).

Over the first stage of RI (RI < 26% from day 18001-21899 in simulation), the SIAs-based strategies (either the RI + SIAs strategy or the “switch” strategies) result in the lowest accumulation of cases, as expected from Figure 3. Note that PIRIs in Stage 1, which are implemented in the two scenarios using the PIRI-based strategies, are limited to a maximum rate ratio of 7-times RI (because the actual MSRR is too high). Over Stage 2 of RI (RI between 26% and 70%; day 21900-28499 in simulation), PIRI-based strategies (either RI + PIRI or the “switch” strategies) lead to lower accumulation of cases. This holds for both PIRIs conducted at a constant RR of 7 or at the MSRR. Even though SIAs are optimal at high levels of RI in Stage 3 (suggested by Figure 3), the number of cases in the “switch” and “RI+SIAs” scenarios were only marginally smaller than the “RI+PIRI” scenarios in Stage 3. This arises despite our limitation that PIRIs in Stage 3 were limited to a RR of only 7-times RI. Thus, while SIAs may be technically optimal at constant high RI, the difference between PIRI and SIAs-based strategies is small once RI is high. In all PIRI-based scenarios, cumulative caseloads by day 33,000 were lower than in the “RI+SIAs” strategy.

In Stages 2 and 3, SIA-based strategies (including the RI + SIAs strategy and the “switch” strategies) exhibit minimal case accumulation immediately following the SIAs. However, these periods are followed by sharp surges in incidence characteristic of large-scale outbreaks (Supplemental Figure S6). Conversely, PIRI strategies maintain more consistent case numbers over time with more smooth trajectories, suggesting they are more effective at preventing these substantial outbreaks caused by diseases resurgence.

## Discussion

We found that annual PIRIs with vaccination rate higher than a minimally sufficient multiplier of the daily RI rate can result in fewer cases than quinquennial SIAs with 70% coverage over a broad range of RI coverage levels and demographic settings. Specifically, even selective (only targeting unvaccinated children) PIRIs conducted annually result in slower accumulation of the susceptible and lower incidence than the larger and non-selective SIAs conducted at larger intervals. We characterized the operational performance of PIRI-based strategies in terms of PIRI’s rate of vaccination (number of vaccinations per unvaccinated child per day) relative to RI and showed that the minimally sufficient rate ratio required to exceed the performance of SIAs depends on RI coverage, birth rate, SIAs characteristics, PIRI’s selectivity (Figure 1). Identifying this minimally sufficient rate can guide program transition from SIAs to PIRIs based on the local programmatic conditions.

The minimally sufficient rate ratio (MSRR) of PIRI is a function of the level of RI coverage. In scenarios where RI coverage is low, a temporary, annual increase in the vaccination rate via PIRI (e.g., for one month) within a restricted age window (9–24 or 9–36 months) would need to be substantially higher (possibly infinite) to reduce case numbers below those achieved by an SIAs-based strategy. Note that by comparison, an SIAs necessarily achieve a very high daily rate of vaccination to achieve high coverage of the target population in a short window. In scenarios where the RI coverage is very high, the majority of children would have been vaccinated by RI, and children remaining susceptible are more likely to be the results of primary vaccine failures. As strictly defined, PIRI is only aimed at giving the unvaccinated children their first dose of vaccine, thus not addressing this source of susceptibility; on the contrary, non-selective SIAs do. However, this doesn’t mean PIRI is necessarily a poor choice of vaccination in the countries with high RI coverage. The performance difference between PIRI and SIAs is marginal when the overall RI coverage level is high (Figure 5). Moreover, switching from selective to non-selective PIRI — e.g. targeting 0-dose kids as well as 1-dose kids — will enhance the performance of PIRI (Figure 4B) and address susceptibility due to primary vaccine failure in high-RI-coverage settings.

**Figure 5.**
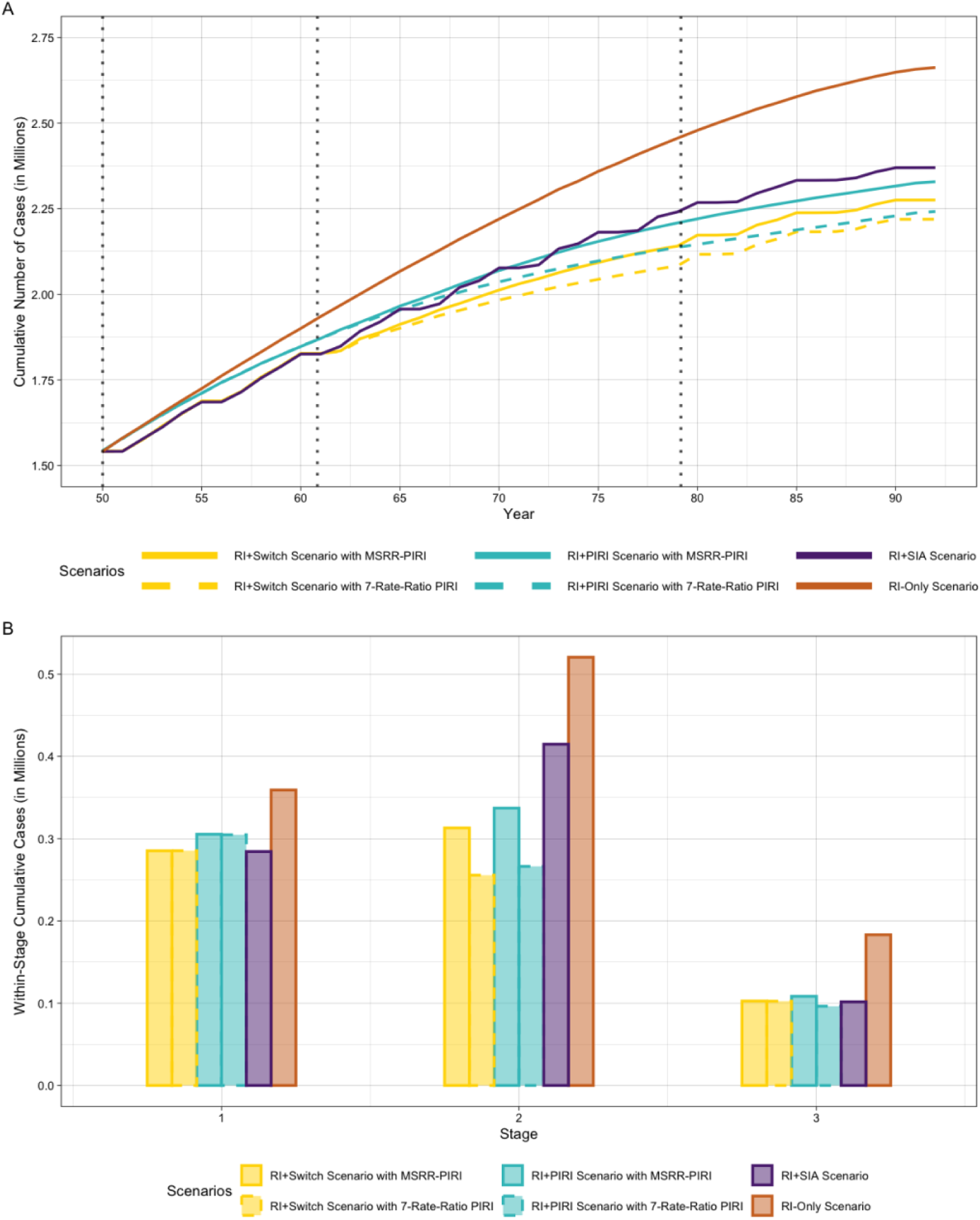
The numbers of measles cases under 6 different scenarios, which are averaged across 500 simulations in each scenario, are cumulatively recorded from day 18,001 to day 33,000 with RI coverage level increasing from 0% to 100%. The whole period is divided into stages from 1 to 3, corresponding to the settings with low (<26%), intermediate (between 26% and 70%), and high (>70%) level of RI coverage, respectively. Panel A illustrates the changes in the cumulative cases over time with dash lines separating three stages. Panel B illustrates the sums of the cases within each stage.

We have chosen to frame the discussion of PIRI in terms of rates, and MSRR, instead of coverage for several reasons. First, the vaccination rate is an observable and measurable metric in real time, despite the differences in time scales between RI (continuous) and PIRI (pulsed). The number of persons getting vaccinated at a specific clinic per day is, in principle, recordable and can be used as a good approximation of the vaccination rate. Vaccination coverage has been a standard indicator of program performance ^41,42^, but is often available only infrequently because of the time and effort to measure. Because they are conducted over short intervals of time, the coverage of PIRIs and SIAs, can be estimated by representative sampling and community surveys with the population eligible at the time of the campaign as the sampling frame ^43^; the population eligible for a selective PIRI (i.e. those within the age range and previously vaccinated) is itself a frequently unknown quantity. These surveys often add operational costs to the campaign and estimates are available often months after the campaign^44^. This additional cost and delay in assessment may be prohibitive for frequent (i.e. annual) activities. Second, PIRI, by definition, is an intensification of RI, and thus RI is the implied frame of reference. However, estimates of RI coverage derived from a single, even well-designed survey may become rapidly obsolete, given the continuous nature of the RI process. Administrative coverage of RI, though available frequently, is biased often due to uncertainty about the denominator (the eligible population); surveillance-based proxies of coverage have been proposed but are not yet operationally validated ^45^. Rates, measured as vaccinations per unit time of the activity, allow for direct comparison between RI and PIRI and serves as a common metric despite the time-scale differences of the activities. Third, the rate ratio between PIRI and RI directly reflects the additional amount of effort needed to intensify the existing RI, which aligns with the definition of PIRI. For example, a MSRR at 3 means during the PIRI month, at least 3 times the amounts of vaccines in a non-PIRI month must be administered to outperform the status quo SIAs program. This rate ratio sets a very clear, quantifiable, and executable goal for the PIRI vaccination program.

This study provides a framework for comparisons between PIRI and SIAs to support transitions away from vaccination strategies that have relied heavily on SIAs ^6^. While this analysis cannot be exhaustive of all PIRI or SIA strategies, nor specific to every operational setting, it delineates the essential operational characteristics of PIRIs and provides a mathematical framework for comparing the relative performance of mixed delivery strategies (e.g., RI plus PIRI and RI plus SIAs) to guide program planning. The status quo of measles vaccination programs is “RI+SIAs” and it’s faced with multiple challenges. The rate of improvement in RI coverage has slowed, or stalled, in many countries for years ^46^. SIAs are very expensive to carry out ^47^ and very often rely on external funding sources like Gavi ^48^. In practice, the administration cost of SIAs can be very unpredictable and may exceed the budget ^48^. Integration of different vaccination programs is sometimes put in place in the face of limited budget ^49^, which might overstretch the capacities of local health facilities and compromise the effects of campaigns. Furthermore, “campaign fatigue” was expressed in communities that have gone through repeated SIAs, and this contributed to vaccine hesitancy and the spread of misinformation ^50^.

PIRI has potential advantages over SIAs. First, PIRI conducted at higher frequency than SIAs shorten the durations at which children missed by RI may be exposed to transmission (e.g. from up to 5 years to up to 1 year in our simulations). Reaching zero-dose children is a primary objective of both SIAs and PIRI. While the efficacy of each intervention varies ^15^, evidence indicates that both SIAs and PIRI successfully reach children missed by RI ^26,28,51^.

A WHO program report on PIRI’s impact in Bosnia and Herzegovina found that PIRI effectively improved the Measles, Mumps, and Rubella (MMR) vaccine coverage from roughly 69% to 76% over a 10-month period ^52^. Second, PIRI may be more financially advantageous than SIAs since it leverages the existing RI infrastructure rather than establishing parallel delivery systems. While large SIAs may achieve low per-unit cost due to its scale effect ^19^, recent evaluations of large-scale PIRI initiatives, such as India’s Intensified Mission Indradhanush, demonstrated that PIRI remains cost-effective ($83 per zero-dose child) ^19^. Unlike SIAs, which require repeated infusions of emergency ‘surge’ funding ^47–49^, PIRI investments contribute to the operational capacity of local health centers, potentially reducing the marginal cost of reaching under-vaccinated populations over time ^24^. Third, PIRI not only increases vaccine coverage in the short term but may also sustains secondary gains over the long term ^53^. Beyond the catch-up in vaccination coverage, PIRI cultivates long-term system resilience ^54^ by strengthening coordination channels and normalizing health-seeking behavior in underserved communities ^29,52^. Even in contexts where post-campaign retention wanes, these intensifications serve as valuable diagnostic stress tests, highlighting the specific structural barriers that must be addressed to convert temporary awareness into sustained engagement ^27^. This is partially because PIRI is conducted with more consistent vaccination effort and operates as a part of RI to improve the overall vaccine accessibility.

PIRI is also subject to some concerns regarding its impacts or feasibilities. First, the factors impeding improvements in RI may also affect the performance of PIRI ^30^. Health workers exposed to higher financial incentives associated with PIRI, might be reluctant to be fully committed to RI services ^25,55^, which might hinder improvements in RI in the long term. PIRI is not always efficient if not optimized. The 2017-2018 national PIRI in India was only estimated to increase infant vaccines by 0.9 to 2.2 doses per PIRI service session ^27^. It’s also important to consider that PIRI may not be feasible to carry out annually or semi-annually in areas with weaker health systems, where the comparably less frequent SIAs may be deeply integrated, more cost-effective, and too essential for outbreak prevention to replace ^28^.

In practice, PIRI programs exhibit significant operational heterogeneity and may require substantial design optimization. We create this quantitative framework to delineate PIRI’s essential characteristics to facilitate the communication and evaluation of PIRI programs. Specifically, we utilized vaccination rates of PIRI relative to RI instead of absolute coverage. Through this framework, we can address concrete designs of PIRI and identify suitable scenarios for future implementations. Notably, a cost-effectiveness analysis is excluded from this study; we assume that establishing PIRI’s operational feasibility and potential to substitute for SIAs is a prerequisite to economic evaluation. The assessments of PIRI’s cost-effectiveness are preserved for the future after costs and dose requirements associated with SIAs and PIRI become more available.

Here we showed that PIRI may be a bridge to transition away from SIAs while emphasizing the need for continuous investments into RI improvements. The mathematical framework here is a starting point for further formalization of PIRI models to support vaccination program designs.

## Data and Code Availability

All the raw data generated in this study will be available upon request. The code will be accessible on GitHub (currently in the process to be organized and submitted).

## Acknowledgements

We would like to thank all the members from the Ferrari Lab and everyone else who made constructive suggestions for their support and contribution.

## References

1. Minta, A. A. Progress Toward Measles Elimination — Worldwide, 2000–2023. MMWR Morb Mortal Wkly Rep 73, (2024).

2. Clements, C. J. et al. Researching routine immunization–do we know what we don’t know? Vaccine 29, 8477–8482 (2011).

3. Fields, R., Dabbagh, A., Jain, M. & Sagar, K. S. Moving forward with strengthening routine immunization delivery as part of measles and rubella elimination activities. Vaccine 31 Suppl 2, B115–121 (2013).

4. Wallace, A. S. et al. Impact of an Intervention to Use a Measles, Rubella, and Polio Mass Vaccination Campaign to Strengthen Routine Immunization Services in Nepal. J Infect Dis 216, S280–S286 (2017).

5. Portnoy, A., Jit, M., Helleringer, S. & Verguet, S. Impact of measles supplementary immunization activities on reaching children missed by routine programs. Vaccine 36, 170–178 (2018).

6. Cutts, F. T., Ferrari, M. J., Krause, L. K., Tatem, A. J. & Mosser, J. F. Vaccination strategies for measles control and elimination: time to strengthen local initiatives. BMC Medicine 19, 2 (2021).

7. Orenstein, W. A., Hinman, A., Nkowane, B., Olive, J. M. & Reingold, A. Measles and Rubella Global Strategic Plan 2012-2020 midterm review. Vaccine 36 Suppl 1, A1–A34 (2018).

8. Measles vaccines: WHO position paper – April 2017. Wkly Epidemiol Rec 92, 205–227 (2017).

9. McKee, A., Ferrari, M. J. & Shea, K. The effects of maternal immunity and age structure on population immunity to measles. Theor Ecol 8, 261–271 (2015).

10. Adebisi, Y. A. & Lucero-Prisno, D. E. Fixing Data Gaps for Population Health in Africa: An Urgent Need. Int J Public Health 67, 1605418 (2022).

11. Umeh, G. C., et al. Micro-planning for immunization in Kaduna State, Nigeria: Lessons learnt, 2017. Vaccine 36, 7361–7368 (2018).

12. Clements, C. J., Soakai, T. S. & Sadr-Azodi, N. A review of measles supplementary immunization activities and the implications for Pacific Island countries and territories. Expert Rev Vaccines 16, 161–174 (2017).

13. Cutts, F. T. et al. Using models to shape measles control and elimination strategies in low- and middle-income countries: A review of recent applications. Vaccine 38, 979–992 (2020).

14. Goodson, J. L., Alexander, J. P., Linkins, R. W. & Orenstein, W. A. Measles and rubella elimination: learning from polio eradication and moving forward with a diagonal approach. Expert Rev Vaccines 16, 1203–1216 (2017).

15. Prosperi, C. et al. Added value of the measles-rubella supplementary immunization activity in reaching unvaccinated and under-vaccinated children, a cross-sectional study in five Indian districts, 2018-20. Vaccine 41, 486–495 (2023).

16. Yang, Y. et al. Challenges Addressing Inequalities in Measles Vaccine Coverage in Zambia through a Measles–Rubella Supplementary Immunization Activity during the COVID-19 Pandemic. Vaccines 11, 608 (2023).

17. Vaughan, K. et al. The costs of delivering vaccines in low- and middle-income countries: Findings from a systematic review. Vaccine: X 2, 100034 (2019).

18. Graham, M. et al. Measles and the canonical path to elimination. Science 364, 584–587 (2019).

19. Kaucley, L. & Levy, P. Cost-effectiveness analysis of routine immunization and supplementary immunization activity for measles in a health district of Benin. Cost Effectiveness and Resource Allocation 13, 14 (2015).

20. Prosperi, C. et al. Added value of the measles-rubella supplementary immunization activity in reaching unvaccinated and under-vaccinated children, a cross-sectional study in five Indian districts, 2018-20. Vaccine 41, 486–495 (2023).

21. Verguet, S. et al. Impact of supplemental immunisation activity (SIA) campaigns on health systems: findings from South Africa. J Epidemiol Community Health 67, 947–952 (2013).

22. Hagan, J. et al. Challenges for Sustaining Measles Elimination: Post-Verification Large-Scale Import-Related Measles Outbreaks in Mongolia and Cambodia, Resulting in the Loss of Measles Elimination Status. Vaccines (Basel) 12, 821 (2024).

23. Winter, A. K. et al. Feasibility of measles and rubella vaccination programmes for disease elimination: a modelling study. Lancet Glob Health 10, e1412–e1422 (2022).

24. IMMUNIZATIONbasics Project, U.S. Agency for International Development, & World Health Organization. Periodic Intensification of ROUTINE IMMUNIZATION--Lessons Learned and Implications for Action. (2009).

25. Ryman, T. K., Trakroo, A., Ekka, J. B. & Watkins, M. Contribution of Immunization Weeks toward improving coverage, access to services, and completion of recommended childhood vaccinations in Assam, India. Vaccine 30, 2551–2555 (2012).

26. Clarke-Deelder, E. et al. Health impact and cost-effectiveness of expanding routine immunization coverage in India through Intensified Mission Indradhanush. Health Policy Plan 39, 583–592 (2024).

27. Clarke-Deelder, E. et al. Impact of campaign-style delivery of routine vaccines: a quasi-experimental evaluation using routine health services data in India. Health Policy Plan 36, 454–463 (2021).

28. Oliphant, N. P. et al. The Contribution of Child Health Days to Improving Coverage of Periodic Interventions in Six African Countries. Food Nutr Bull 31, S248–S263 (2010).

29. Mihigo, R. et al. African vaccination week as a vehicle for integrated health service delivery. BMC Health Serv Res 15, 358 (2015).

30. Summan, A., Nandi, A., Deo, S. & Laxminarayan, R. Improving vaccination coverage and timeliness through periodic intensification of routine immunization: evidence from Mission Indradhanush. Ann N Y Acad Sci 1502, 110–120 (2021).

31. Gurnani, V. et al. Improving vaccination coverage in India: lessons from Intensified Mission Indradhanush, a cross-sectoral systems strengthening strategy. BMJ 363, k4782 (2018).

32. Njoh, A. A. et al. Impact of periodic intensification of routine immunization within an armed conflict setting and COVID-19 outbreak in Cameroon in 2020. Confl Health 16, 29 (2022).

33. Gillespie, D. T. Approximate accelerated stochastic simulation of chemically reacting systems. J. Chem. Phys. 115, 1716–1733 (2001).

34. Schenzle, D. An age-structured model of pre- and post-vaccination measles transmission. IMA J Math Appl Med Biol 1, 169–191 (1984).

35. Guerra, F. M. et al. The basic reproduction number (R0) of measles: a systematic review. The Lancet Infectious Diseases 17, e420–e428 (2017).

36. Keeling, M. J. & Rohani, P. Stochastic Dynamics. in Modeling Infectious Diseases in Humans and Animals 190–231 (Princeton University Press, 2008). doi:10.2307/j.ctvcm4gk0.9.

37. Papadopoulos, T., Jit, M., Ferrari, M. J. & Vynnycky, E. Factors determining the overlap between recipients of the first and second dose of measles vaccine in nineteen surveys. Sci Rep 15, 30737 (2025).

38. Carazo, S., Billard, M.-N., Boutin, A. & De Serres, G. Effect of age at vaccination on the measles vaccine effectiveness and immunogenicity: systematic review and meta-analysis. BMC Infect Dis 20, 251 (2020).

39. Lessler, J., Metcalf, C. J. E., Cutts, F. T. & Grenfell, B. T. Impact on Epidemic Measles of Vaccination Campaigns Triggered by Disease Outbreaks or Serosurveys: A Modeling Study. PLOS Medicine 13, e1002144 (2016).

40. Thakkar, N., Gilani, S. S. A., Hasan, Q. & McCarthy, K. A. Decreasing measles burden by optimizing campaign timing. Proceedings of the National Academy of Sciences 116, 11069–11073 (2019).

41. Patel, C. et al. Measuring National Immunization System Performance: A Systematic Assessment of Available Resources. Glob Health Sci Pract 11, e220055 (2023).

42. Galles, N. C. et al. Measuring routine childhood vaccination coverage in 204 countries and territories, 1980–2019: a systematic analysis for the Global Burden of Disease Study 2020, Release 1. The Lancet 398, 503–521 (2021).

43. Danovaro-Holliday, M. C., Koh, M., Steulet, C., Rhoda, D. A. & Trimner, M. K. Lessons from Recent Measles Post-Campaign Coverage Surveys Worldwide. Vaccines (Basel) 12, 1257 (2024).

44. Cutts, F. T., Danovaro-Holliday, M. C. & Rhoda, D. A. Challenges in measuring supplemental immunization activity coverage among measles zero-dose children. Vaccine 39, 1359–1363 (2021).

45. Bhatia, D. et al. Prediction of subnational-level vaccination coverage estimates using routine surveillance data and survey data. Vaccine 60, 127277 (2025).

46. Haeuser, E. et al. Global, regional, and national trends in routine childhood vaccination coverage from 1980 to 2023 with forecasts to 2030: a systematic analysis for the Global Burden of Disease Study 2023. The Lancet 406, 235–260 (2025).

47. Sibeudu, F. T., Onwujekwe, O. E. & Okoronkwo, I. L. Cost analysis of supplemental immunization activities to deliver measles immunization to children in Anambra state, south-east Nigeria. Vaccine 38, 5947–5954 (2020).

48. Boonstoppel, L. et al. Cost of integrated immunization campaigns in Nigeria and Sierra Leone: bottom-up costing studies. BMC Health Services Research 24, 1334 (2024).

49. Jean Baptiste, A. E., et al. The cost of implementing measles campaign in Nigeria: comparing the stand-alone and the integrated strategy. Health Econ Rev 13, 36 (2023).

50. Krishnendhu, V. K. & George, L. S. Drivers and barriers for measles rubella vaccination campaign: A qualitative study. Journal of Family Medicine and Primary Care 8, 881 (2019).

51. Farahani, M. et al. Evaluation of Integrated Child Health Days as a Catch-Up Strategy for Immunization in Three Districts in Uganda. Vaccines (Basel) 12, 1353 (2024).

52. World Health Organization. Periodic intensification of routine immunization activities increase vaccination in Bosnia and Herzegovina. https://www.who.int/about/accountability/results/who-results-report-2020-mtr/country-story/2023/periodic-intensification-of-routine-immunization-activities-increase-vaccination-in-bosnia-and-herzegovina (2024).

53. Njoh, A. A. et al. Impact of periodic intensification of routine immunization within an armed conflict setting and COVID-19 outbreak in Cameroon in 2020. Confl Health 16, 29 (2022).

54. Jones, C. E. Routine Vaccination Coverage — Worldwide, 2023. MMWR Morb Mortal Wkly Rep 73, (2024).

55. Doherty, T. et al. Moving from vertical to integrated child health programmes: experiences from a multi-country assessment of the Child Health Days approach in Africa. Tropical Medicine & International Health 15, 296–305 (2010).

