## Supplement for "Periodic intensification of routine immunization (PIRI): modeling a novel strategy to supplement routine and pulsed measles vaccination"

Supplement/Extended Data

SFigure 1. The minimally sufficient rate ratio (MSRR) between PIRI and RI to achieve equivalent cases as SIA at different levels of RI coverage during the period of stable dynamics (from day 5001 to day 10,000). PIRI targets kids aged 6 to 18 months who received zero dose of vaccine from RI before. The RI coverage levels vary from one simulation to another, but it remains unchanged within each scenario. SIA has different intervals or coverages across different scenarios but remains constant within each scenario.

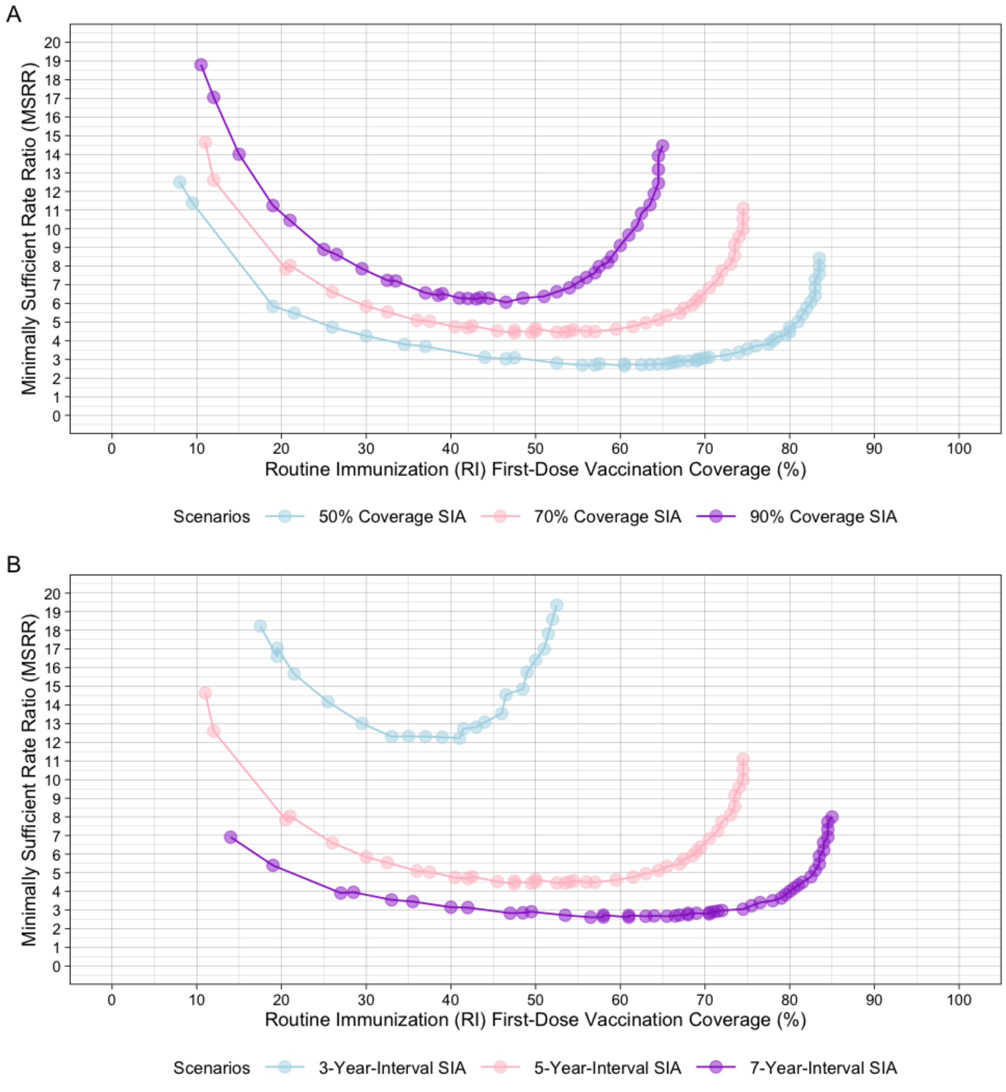

SFigure 2. The sufficient rate ratio between PIRI and RI to achieve equivalent or fewer cases than SIA at different levels of RI coverage during the period of stable dynamics (from day 5001 to day 10,000). PIRI targets kids aged 6 to 18 months who received zero dose of vaccine from RI before. The RI coverage levels vary from one simulation to another, but it remains unchanged within each scenario. SIA always has a fixed 5-year interval and a 70% coverage amongst all kids under 5 across all simulations. PIRI's vaccination rate remains fixed throughout each simulation but differs across different simulations. RI only administers the first-dose vaccines and no second-dose vaccine.

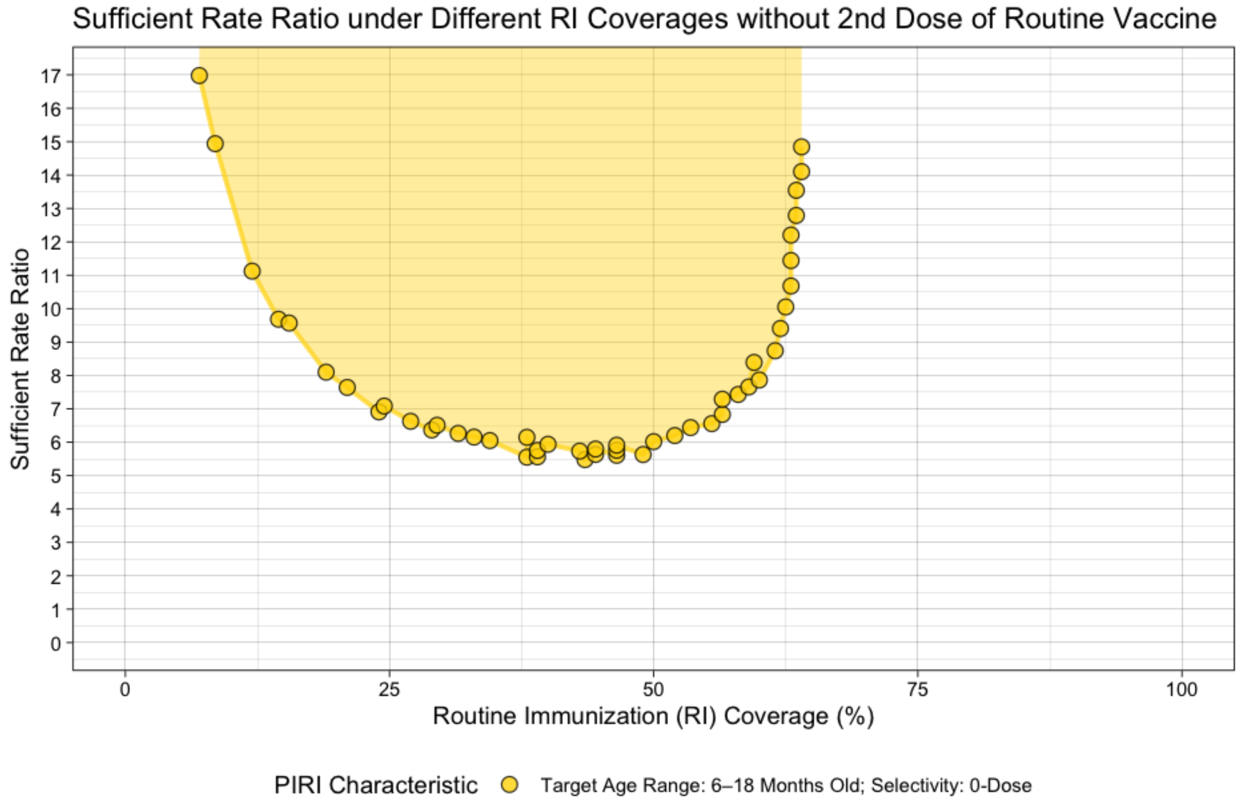

SFigure 3. The sufficient rate ratio between PIRI and RI to achieve equivalent or fewer cases than SIA at different levels of RI coverage during the period of stable dynamics (from day 5001 to day 10,000). PIRI targets kids aged 6 to 18 months who received zero dose of vaccine from RI before. The RI coverage levels vary from one simulation to another, but it remains unchanged within each scenario. SIA always has a fixed 5-year interval and a 70% coverage amongst all kids under 5 across all simulations. PIRI's vaccination rate remains fixed throughout each simulation but differs across different simulations. Different from Figure 3, the rate ratio metric used in this figure is the following: $\frac{\int_{A_{lower}^{PIRI}}^{A_{upper}^{PIRI}}(v_a^{PIRI})\,da}{\int_{A_{lower}^{1}}^{A_{upper}^{1}}(v_a^{1})\,da}$.

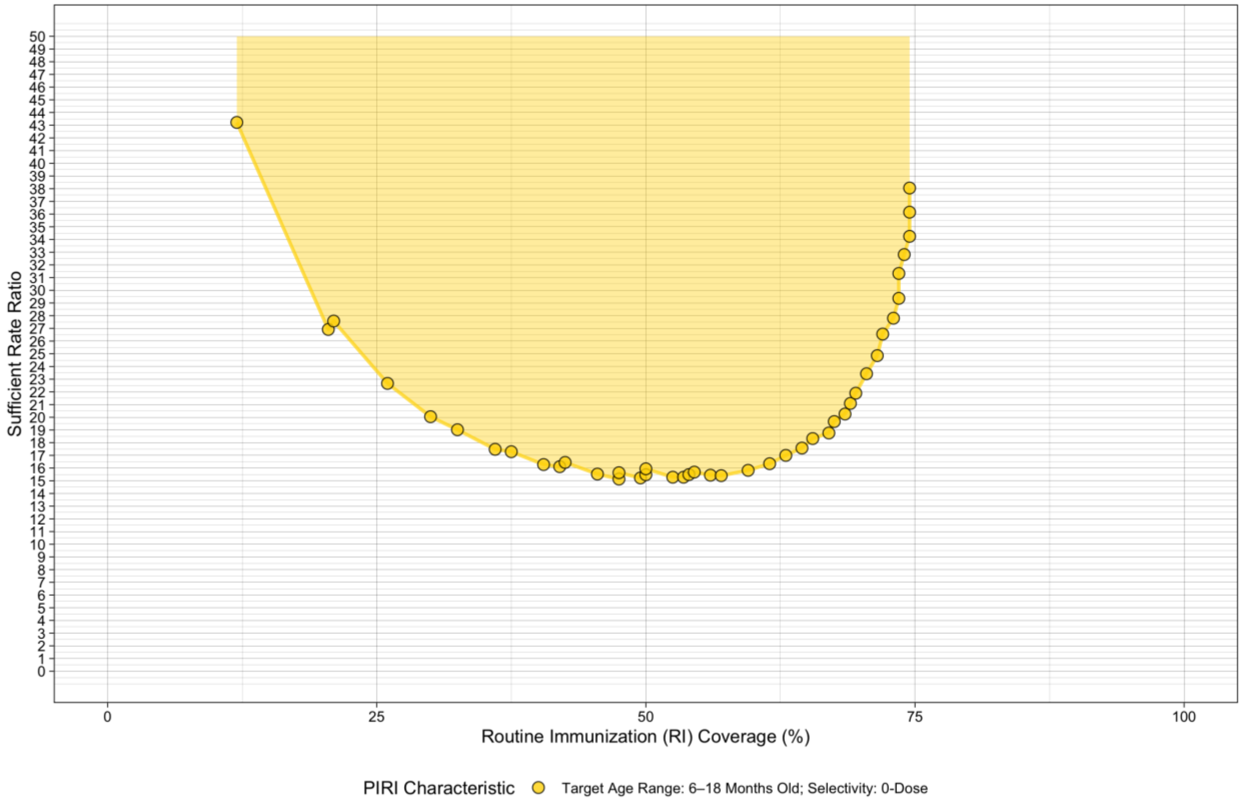

SFigure 4. The minimally sufficient rate ratio (MSRR) across different levels of RI coverage under different assumptions of PIRI characteristics, SIA characteristics, and demographic characteristics. Panel A demonstrates the MSRR curves under PIRIs with different ranges of target age group, from 12-month-long (the default, same as in Figure 3) to 36-month-long. Panel B shows the MSRR curve when PIRI targets only 0-dose kids (default) or 0- and 1-dose kids. Panel C compares MSRR curves when both PIRI and SIA are implemented in a specific month relative to the peak month of the measles transmission, with the default scenario as the one where both SIA and PIRI are implemented 3 months ahead of the peak month. Panel D illustrates MSRR curves given different birth rates in the population, from 30 new births per 1000 people at reproductive ages per year to 70 with the 50 as the default. Different from Figure 4, the rate ratio metric used in this figure is the following:

$$\frac{\int_{A_{lower}^{PIRI}}^{A_{upper}^{PIRI}} (\nu_a^{PIRI})\, da}{\int_{A_{lower}^{1}}^{A_{upper}^{1}} (\nu_a^{1})\, da}.$$

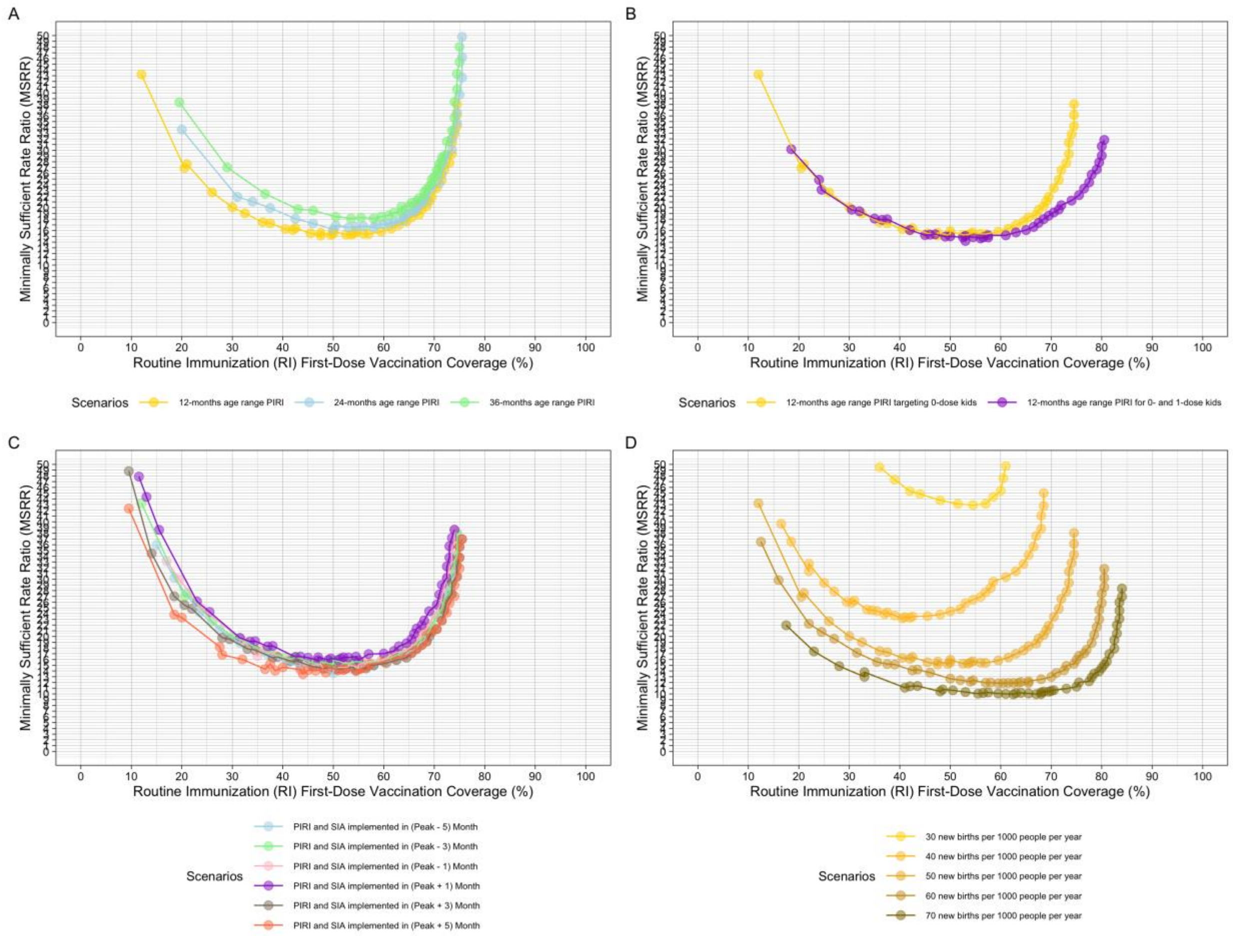

SFigure 5. The minimally sufficient rate ratio (MSRR) across different levels of RI coverage under different assumptions of PIRI characteristics, SIA characteristics, and demographic characteristics. Panel A demonstrates the MSRR curves under PIRIs with different ranges of target age group, from 12-month-long (the default, same as in Figure 3) to 36-month-long. Panel B shows the MSRR curve when PIRI targets only 0-dose kids (default) or 0- and 1-dose kids. Panel C compares MSRR curves when both PIRI and SIA are implemented in a specific month relative to the peak month of the measles transmission, with the default scenario as the one where both SIA and PIRI are implemented 3 months ahead of the peak month. Panel D illustrates MSRR curves given different birth rates in the population, from 30 new births per 1000 people at reproductive ages per year to 70 with the 50 as the default. The rate ratio metric used in this figure is the same as that used in Figure 4.

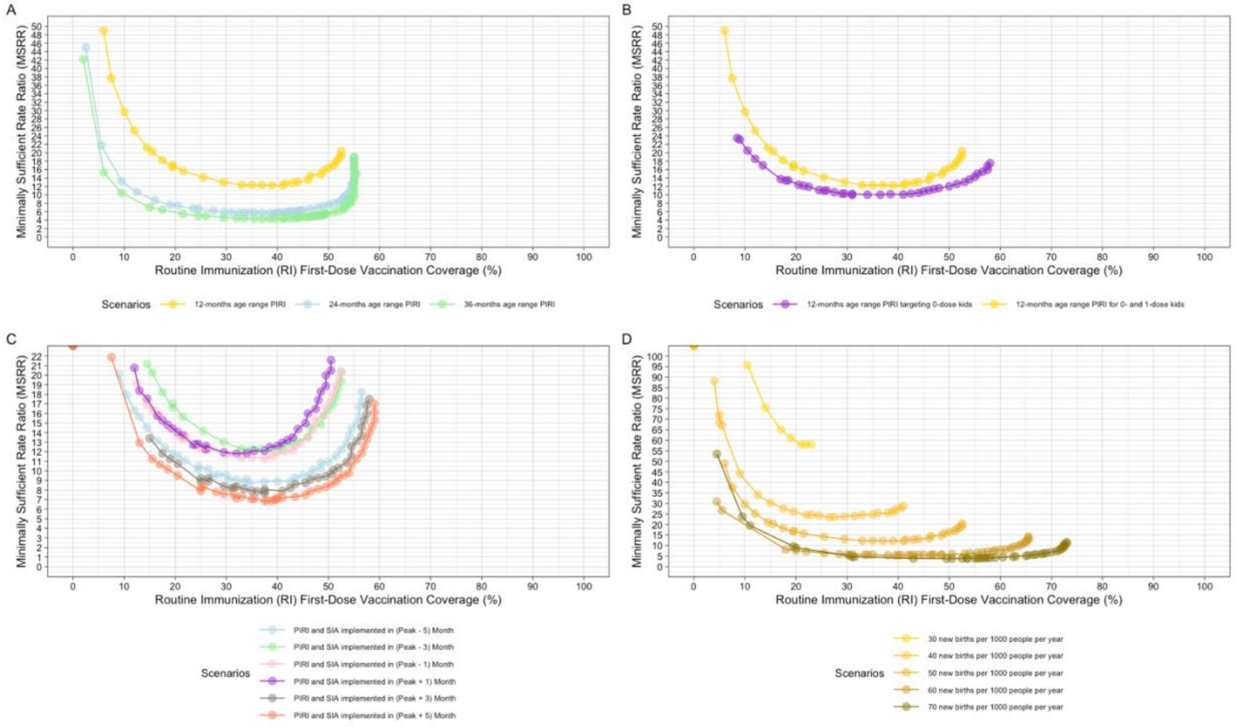

SFigure 6. The annual numbers of measles cases under 2 different scenarios—RI plus SIAs across all three stages (in purple) and RI plus PIRI with a fixed rate ratio across three stages (in green). The middle lines are the case means averaged across 500 simulations in each scenario; the lower and upper bounds represent the 5th and 95th percentile of the annual cases across the 500 simulations, respectively; and the shaded area represents the variability in the annual cases among the simulations between the 5th and 95th percentiles. The whole period is divided into stages from 1 to 3, corresponding to the settings with low (<26%), intermediate (between 26% and 70%), and high (>70%) level of RI coverage, respectively.

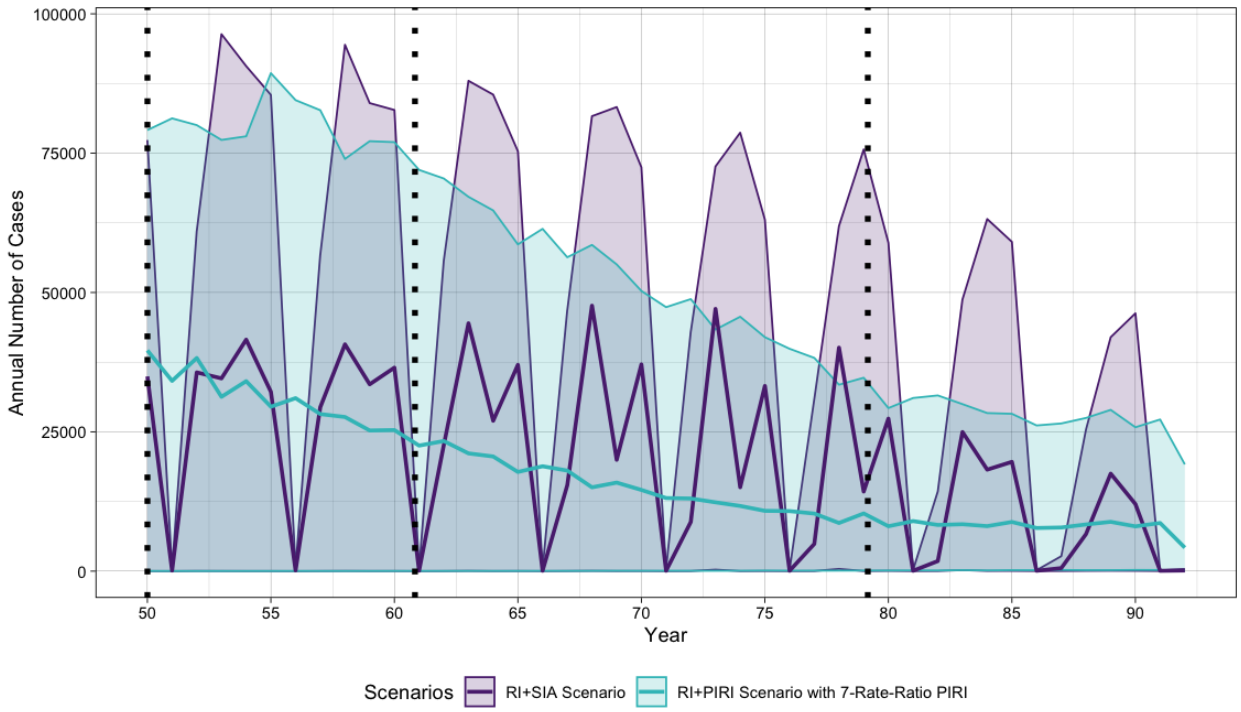

STable 1. Initial overall population size in each age group

| Age Group | Initial Population Size (number of people) |
|---|---|
| New birth | 1997 |
| 1 month old | 1995 |
| 2 months old | 1993 |
| 3 months old | 1991 |
| 4 months old | 1989 |
| 5 months old | 1987 |
| 6 months old | 1985 |
| 7 months old | 1983 |
| 8 months old | 1980 |
| 9 months old | 1978 |
| 10 months old | 1976 |
| 11 months old | 1974 |
| 12 months old | 1972 |
| 13 months old | 1970 |
| 14 months old | 1968 |
| 15 months old | 1966 |
| 16 months old | 1964 |
| 17 months old | 1962 |
| 18 months old | 1960 |
| 19 months old | 1958 |
| 20 months old | 1956 |
| 21 months old | 1953 |
| 22 months old | 1951 |
| 23 months old | 1949 |
| 24 months old | 1947 |
| 25 months old | 1945 |
| 26 months old | 1943 |
| 27 months old | 1941 |
| 28 months old | 1939 |
| 29 months old | 1937 |
| 30 months old | 1935 |
| 31 months old | 1933 |
| 32 months old | 1931 |
| 33 months old | 1928 |
| 34 months old | 1926 |

| | |
|---|---|
| 35 months old | 1924 |
| 36 months old | 1922 |
| 37 months old | 1920 |
| 38 months old | 1918 |
| 39 months old | 1916 |
| 40 months old | 1914 |
| 41 months old | 1912 |
| 42 months old | 1910 |
| 43 months old | 1908 |
| 44 months old | 1906 |
| 45 months old | 1903 |
| 46 months old | 1901 |
| 47 months old | 1899 |
| 48 months old | 1897 |
| 49 months old | 1895 |
| 50 months old | 1893 |
| 51 months old | 1891 |
| 52 months old | 1889 |
| 53 months old | 1887 |
| 54 months old | 1885 |
| 55 months old | 1883 |
| 56 months old | 1881 |
| 57 months old | 1878 |
| 58 months old | 1876 |
| 59 months old | 1874 |
| 60 months old | 1872 |
| 6 years old | 22330 |
| 7 years old | 22030 |
| 8 years old | 21730 |
| 9 years old | 21431 |
| 10 years old | 21131 |
| 11 years old | 20831 |
| 12 years old | 20531 |
| 13 years old | 20232 |
| 14 years old | 19932 |
| 15 years old | 19632 |
| 16 years old | 19332 |
| 17 years old | 19033 |
| 18 years old | 18733 |

| | |
|---|---|
| 19 years old | 18433 |
| 20 years old | 18134 |
| 21 years old | 17834 |
| 22 years old | 17534 |
| 23 years old | 17234 |
| 24 years old | 16935 |
| 25 years old | 16635 |
| 26 years old | 16335 |
| 27 years old | 16035 |
| 28 years old | 15736 |
| 29 years old | 15436 |
| 30 years old | 15136 |
| 31 years old | 14837 |
| 32 years old | 14537 |
| 33 years old | 14237 |
| 34 years old | 13937 |
| 35 years old | 13638 |
| 36 years old | 13338 |
| 37 years old | 13038 |
| 38 years old | 12738 |
| 39 years old | 12439 |
| 40 years old | 12139 |
| 41 years old | 11839 |
| 42 years old | 11540 |
| 43 years old | 11240 |
| 44 years old | 10940 |
| 45 years old | 10640 |
| 46 years old | 10341 |
| 47 years old | 10041 |
| 48 years old | 9741 |
| 49 years old | 9441 |
| 50 years old | 9142 |
| 51 years old | 8842 |
| 52 years old | 8542 |
| 53 years old | 8243 |
| 54 years old | 7943 |
| 55 years old | 7643 |
| 56 years old | 7343 |
| 57 years old | 7044 |

| | |
|---|---|
| 58 years old | 6744 |
| 59 years old | 6444 |
| 60 years old | 6144 |
| 61 years old | 5845 |
| 62 years old | 5545 |
| 63 years old | 5245 |
| 64 years old | 4946 |
| 65 years old | 4646 |
| 66 years old | 4346 |
| 67 years old | 4046 |
| 68 years old | 3747 |
| 69 years old | 3447 |
| 70 years old | 3147 |
| 71 years old | 2847 |
| 72 years old | 2548 |
| 73 years old | 2248 |
| 74 years old | 1948 |
| 75 years old | 1649 |
| 76 years old | 1349 |
| 77 years old | 1049 |
| 78 years old | 749 |
| 79 years old | 450 |
| 80 years old and above | 150 |

STable 2. Age-group-specific birth rates

| Age Groups | Birth Rate |
|---|---|
| Under 18 | 0 |
| Between 18 and 50 | 50.0/1000.0/360.0 |
| 50 and above | 0 |

STable 3. Age-group-specific mortality rates

| Age Group | Mortality Rate |
|---|---|
| New birth | 0.000264123 |
| 1 month old | 0.000114257 |
| 2 months old | 8.77557E-05 |
| 3 months old | 7.40514E-05 |
| 4 months old | 6.53022E-05 |

| | |
|---|---|
| 5 months old | 5.90935E-05 |
| 6 months old | 5.43934E-05 |
| 7 months old | 0.000050676 |
| 8 months old | 0.000047641 |
| 9 months old | 4.51027E-05 |
| 10 months old | 4.29392E-05 |
| 11 months old | 4.10668E-05 |
| 12 months old | 3.94259E-05 |
| 13 months old | 3.79726E-05 |
| 14 months old | 3.66738E-05 |
| 15 months old | 3.55042E-05 |
| 16 months old | 3.44438E-05 |
| 17 months old | 3.34768E-05 |
| 18 months old | 3.25902E-05 |
| 19 months old | 3.17736E-05 |
| 20 months old | 3.10184E-05 |
| 21 months old | 3.03173E-05 |
| 22 months old | 2.96642E-05 |
| 23 months old | 2.90539E-05 |
| 24 months old | 2.84821E-05 |
| 25 months old | 2.79448E-05 |
| 26 months old | 2.74388E-05 |
| 27 months old | 2.69612E-05 |
| 28 months old | 2.65095E-05 |
| 29 months old | 2.60814E-05 |
| 30 months old | 2.56751E-05 |
| 31 months old | 2.52887E-05 |
| 32 months old | 2.49207E-05 |
| 33 months old | 2.45697E-05 |
| 34 months old | 2.42345E-05 |
| 35 months old | 0.000023914 |
| 36 months old | 2.36071E-05 |
| 37 months old | 2.33129E-05 |
| 38 months old | 2.30306E-05 |
| 39 months old | 2.27594E-05 |
| 40 months old | 2.24986E-05 |
| 41 months old | 2.22477E-05 |
| 42 months old | 2.20059E-05 |
| 43 months old | 2.17728E-05 |

| | |
|---|---|
| 44 months old | 0.000021548 |
| 45 months old | 2.13308E-05 |
| 46 months old | 0.000021121 |
| 47 months old | 2.09181E-05 |
| 48 months old | 2.07217E-05 |
| 49 months old | 2.05316E-05 |
| 50 months old | 2.03474E-05 |
| 51 months old | 2.01688E-05 |
| 52 months old | 1.99956E-05 |
| 53 months old | 1.98275E-05 |
| 54 months old | 1.96643E-05 |
| 55 months old | 1.95057E-05 |
| 56 months old | 1.93516E-05 |
| 57 months old | 1.92017E-05 |
| 58 months old | 1.90559E-05 |
| 59 months old | 1.89139E-05 |
| 5 years old | 1.97738E-05 |
| 6 years old | 1.80931E-05 |
| 7 years old | 1.68421E-05 |
| 8 years old | 1.58709E-05 |
| 9 years old | 1.50939E-05 |
| 10 years old | 1.44584E-05 |
| 11 years old | 1.39298E-05 |
| 12 years old | 1.34846E-05 |
| 13 years old | 1.31058E-05 |
| 14 years old | 1.27813E-05 |
| 15 years old | 1.25018E-05 |
| 16 years old | 1.22602E-05 |
| 17 years old | 0.000012051 |
| 18 years old | 1.18698E-05 |
| 19 years old | 1.17133E-05 |
| 20 years old | 1.15785E-05 |
| 21 years old | 1.14632E-05 |
| 22 years old | 1.13654E-05 |
| 23 years old | 1.12837E-05 |
| 24 years old | 1.12166E-05 |
| 25 years old | 1.11633E-05 |
| 26 years old | 1.11227E-05 |
| 27 years old | 1.10942E-05 |

| | |
|---|---|
| 28 years old | 1.10771E-05 |
| 29 years old | 0.000011071 |
| 30 years old | 1.10755E-05 |
| 31 years old | 1.10902E-05 |
| 32 years old | 1.11151E-05 |
| 33 years old | 1.11498E-05 |
| 34 years old | 1.11945E-05 |
| 35 years old | 0.000011249 |
| 36 years old | 1.13135E-05 |
| 37 years old | 0.000011388 |
| 38 years old | 1.14728E-05 |
| 39 years old | 1.15681E-05 |
| 40 years old | 1.16742E-05 |
| 41 years old | 1.17916E-05 |
| 42 years old | 1.19206E-05 |
| 43 years old | 1.20619E-05 |
| 44 years old | 0.000012216 |
| 45 years old | 1.23838E-05 |
| 46 years old | 0.000012566 |
| 47 years old | 1.27637E-05 |
| 48 years old | 1.29778E-05 |
| 49 years old | 1.32097E-05 |
| 50 years old | 1.34609E-05 |
| 51 years old | 1.37329E-05 |
| 52 years old | 1.40277E-05 |
| 53 years old | 1.43474E-05 |
| 54 years old | 1.46946E-05 |
| 55 years old | 1.50721E-05 |
| 56 years old | 1.54834E-05 |
| 57 years old | 1.59324E-05 |
| 58 years old | 1.64238E-05 |
| 59 years old | 0.000016963 |
| 60 years old | 1.75568E-05 |
| 61 years old | 0.000018213 |
| 62 years old | 1.89411E-05 |
| 63 years old | 0.000019753 |
| 64 years old | 2.06632E-05 |
| 65 years old | 2.16898E-05 |
| 66 years old | 0.000022856 |

| | |
|---|---|
| 67 years old | 2.41915E-05 |
| 68 years old | 2.57353E-05 |
| 69 years old | 2.75398E-05 |
| 70 years old | 2.96768E-05 |
| 71 years old | 3.22474E-05 |
| 72 years old | 3.53992E-05 |
| 73 years old | 3.93566E-05 |
| 74 years old | 0.000044479 |
| 75 years old | 5.13811E-05 |
| 76 years old | 6.12131E-05 |
| 77 years old | 7.64215E-05 |
| 78 years old | 0.000103366 |
| 79 years old | 0.000166191 |
| 80 years old and above | 0.00102801 |
